# Water, sanitation, and hygiene practices and challenges during the COVID-19 pandemic: a cross-sectional study in rural Odisha, India

**DOI:** 10.1101/2021.01.26.21250274

**Authors:** Valerie Bauza, Gloria D. Sclar, Alokananda Bisoyi, Fiona Majorin, Apurva Ghugey, Thomas Clasen

## Abstract

Water, sanitation, and hygiene (WASH) practices emerged as a critical component to controlling and preventing the spread of the COVID-19 pandemic. We conducted 131 semi-structured phone interviews with households in rural Odisha, India to understand behavior changes made in WASH practices as a result of the pandemic and challenges that would prevent best practices. Interviews were conducted from May-July 2020 with 73 heads of household, 37 caregivers of children less than five years old, and 21 members of village water and sanitation committees in villages with community-level piped water and high levels of latrine ownership. The majority of respondents (86%, N=104) reported a change in their handwashing practice due to COVID-19 or the related government lockdown, typically describing an increase in handwashing frequency, more thorough washing method, and/or use of soap. These improved handwashing practices remained in place a few months after the pandemic began and were often described as a new consistent practice after additional daily actions (such as returning home), suggesting new habit formation. Few participants (13%) reported barriers to handwashing. Some respondents also detailed improvements in other WASH behaviors including village-level cleaning of water tanks and/or treatment of piped water (48% of villages), household water treatment and storage (17% of respondents), and household cleaning (41% of respondents). However, there was minimal change in latrine use and child feces management practices as a result of the pandemic. We provide detailed thematic summaries of qualitative responses to allow for richer insights into these WASH behavior changes, or lack thereof, during the pandemic. The results also highlight the importance of ensuring communities have adequate WASH infrastructure to enable the practice of safe behaviors and strengthen resilience during a large-scale health crisis.

## Background

COVID-19 rapidly spread across the world and was declared a global pandemic by the World Health Organization (WHO) on March 11, 2020.^1^ As of January 2021, the WHO reports the disease has already caused over 2 million deaths globally.^2^ COVID-19 is caused by the novel coronavirus SARS-CoV-2, which is transmitted person-to-person primarily through inhalation of respiratory droplets/aerosols and contact with fomites contaminated with the virus from an infected individual.^3^ Many preventative measures have been promoted to reduce the spread of the disease, with particular emphasis on avoiding large crowds and social distancing from others, wearing a face mask around others, frequent handwashing with soap, and avoiding touching one’s face.^4^ To quickly limit the spread of the virus, many countries also implemented lockdowns to restrict movement and limit commercial activities. However, access to household water and sanitation facilities can influence the ability of households to adequately practice many of the preventative measures promoted, including frequent handwashing and staying home when infection is suspected or during strict lockdowns.

Additionally, the SARS-CoV-2 virus has been detected in feces of infected individuals, leading to the possibility that fecal-oral transmission could also play a role in virus transmission, particularly in low- and middle-income countries (LMICs) with high rates of open defecation, ineffective fecal sludge management, and poor access to safe drinking water.^5^ An additional factor that could suggest potential foodborne and waterborne transmission is the report of diarrheal symptoms in some individuals infected with COVID-19. SARS-CoV-2 may have higher stability and longer survival times in diarrheal stools, which could increase risk of environmental transmission along potential fecal-oral routes.^6,7^ Although little evidence is available and this is an active area of research, there is no evidence of confirmed transmission of SARS-CoV-2 through food or waterborne routes to date. However, the WHO highlighted the importance of safe management of human feces and treatment of drinking water as precautions against COVID-19 in their interim guidance related to WASH and COVID-19.^8^ The lidless design of squatting pans of flush toilets in many LMICs, including India, is another factor that could potentially lead to a risk of transmission via virus particles becoming aerosolized during flushing and subsequently inhaled or leading to contamination of surfaces.^9^ Due to the recognized importance of handwashing to reduce transmission, and possible spread via feces, it is clear WASH access and practices are important for reducing the spread of COVID-19.

The objective of this research was to understand the WASH-related practices of rural households and communities in Odisha, India in response to the COVID-19 pandemic. We evaluated WASH behavior changes made as a result of COVID-19 and challenges faced that could affect participant’s ability to comply with recommended WASH preventative measures. We used a mix of qualitative and quantitative questions in phone interviews to capture participants’ experiences at a time when COVID-19 was rapidly spreading throughout Odisha, India. The knowledge from this research can help inform WASH-related guidelines for controlling COVID-19 and improve community resilience to external shocks increasingly faced by marginalized populations.

## Methods

### Study site and sampling frame

We conducted semi-structured phone interviews with head of households (HOH), caregivers of children less than 5 years old, and Village Water and Sanitation Committee (VWSC) members in Ganjam and Gajapati districts of rural Odisha, India. Interviews were completed between May-July 2020 when there were lockdowns in both districts, including restrictions on travel and commercial activities which changed over time based on local case counts.^10^ During this time, there was also an influx of migrants workers returning to Odisha, which was accompanied by a surge in local COVID-19 cases and Ganjam district becoming a hotspot of cases within Odisha.^11^ Massive awareness campaigns to educate the public on COVID-19 and encourage preventive measures such as social distancing, mask wearing, and hand hygiene were also underway, with messaging spread through a variety of ways including television, radio, social media, and women’s self-help and other community groups.^12,13^ Details about participants’ knowledge of COVID-19 and impacts on daily life have been previously reported.^11^ We specifically targeted HOHs to understand household-level impacts, caregivers to understand childcare practices including child feces management (CFM), and VWSC members to understand village-level response and changes to village’s piped water system.

All included villages had previously participated in a village-level WASH infrastructure intervention known as MANTRA (Movement and Action Network for Transformation in Rural Areas), that was implemented by the NGO Gram Vikas. This intervention included village-level mobilization for all households to construct a latrine with attached bathing room, and construction of a community piped water system that was connected to households after village-wide latrine construction was completed.^14^ The villages are also currently enrolled in a CFM intervention trial (ISRCTN15831099), but the intervention had not yet started at the time of this study.

### Interview tool

A semi-structured interview tool that included a mix of structured and open-ended questions was developed for each respondent type and has been previously published.^11^ All respondents were asked questions about changes in handwashing, drinking water, sanitation, and household cleaning practices as a result of the pandemic. Respondents were also asked about any challenges they faced with water availability, or challenges in being able to frequently wash their hands with soap. The VWSC interview tool also included questions about changes to village piped water distribution services. Lastly, the caregiver interview tool included specific questions on the impact of the pandemic on CFM practices.

### Data collection

Interviews were conducted by a team of three research assistants, who were all fluent in the local language Odia and originally from Ganjam district. The team underwent a multi-day remote training on how to conduct the phone interviews in May 2020, including training on different interview techniques to use for structured versus open-ended questions. The team piloted the interview tool to gain additional training and to ensure proper translation and comprehension of questions before beginning data collection.

The phone numbers of HOH and caregiver target respondents were randomly ordered using a computer-generated sequence, and research assistants were instructed to contact respondents in the given random order. VWSC members were selected based on village selection. Further details about respondent selection and calling procedures are reported elsewhere.^11^

Data collection occurred over phone calls. When a respondent answered the phone, the research assistants introduced themselves, briefly explained the purpose of the call, and read a consent form, including consent to audio record the conversation. Once the respondent gave their consent, the research assistant continued with the interview questions. During the interview, the research assistant recorded responses and notes in a Microsoft Word version of the interview tool. After the call, research assistants listened to the audio recording to check their transcribed responses and add any details missed. Each interview took approximately 30-45 minutes.

### Data analysis

Stata 16.1 (StataCorp LLC, College Station, Texas, USA) was used for all quantitative analysis of structured survey questions. A modified form of thematic analysis was used for qualitative analysis of open-ended questions and explanations respondents gave about their responses to structured questions. The modified thematic analysis included three rounds of reading through responses to a given question, with memo notes about initial reflections done in the first round, preliminary emergent themes constructed in the second round, and themes finalized in the third round.

During data quality checks, it was realized that one enumerator had sometimes skipped certain survey questions and fabricated some of the data for a portion of their surveys. However, since interviews were audio recorded, the accuracy of the data was ensured by having the study supervisor listen to each of this enumerator’s interviews and correct the data accordingly prior to analysis. As a result, sample sizes vary slightly among different questions due to some questions being sometimes skipped.

### Ethics

Informed consent was verbally obtained over the phone from all participants before interviews began. At the end of the interview, research assistants read a list of helplines participants could call if they needed assistance and also provided information on COVID-19 preventative measures. The study was approved by the Institutional Review Board (IRB) of Emory University (IRB00115339).

## Results

### Characteristics of study participants

In total, 131 participants were interviewed, including 73 heads of household, 21 VWSC members, and 37 caregivers of children <5 years. These respondents spanned 43 villages, including 26 in Ganjam and 17 in Gajapati. Approximately 8% of respondents ended the interviews early, including five HOHs, one VWSC member, and five caregivers. Overall, respondents had high levels of access to household piped water, latrines, and bathing rooms (Table 1). HOH and VWSC participants were predominantly male whereas caregivers were predominantly female (with 88%, 86%, and 8% of each type of respondent being male, respectively).

**Table 1.**
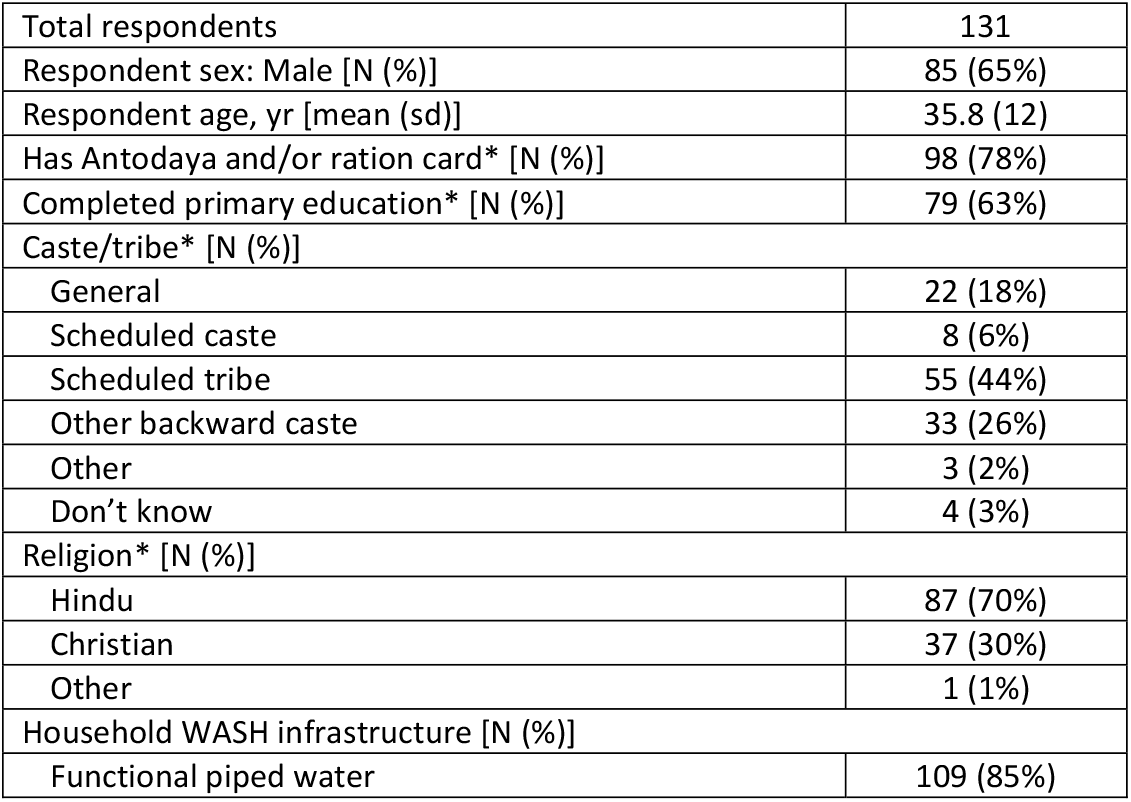
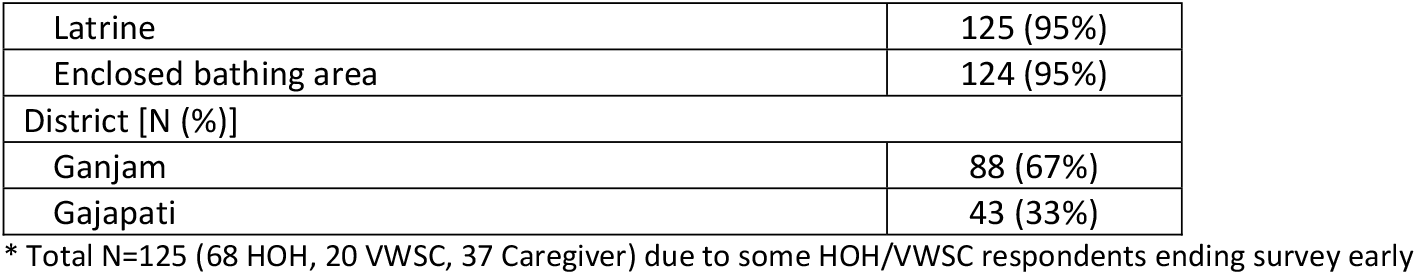
Respondent demographics.

### Handwashing

A large majority of respondents (86%, N=104) reported a change in their handwashing due to COVID-19 or the lockdown. Most described positive changes, such as an increase in the frequency of handwashing, an increase in the use of soap and/or sanitizer, or adopting a more thorough handwashing technique. Several respondents also described that they had started using waterless hand sanitizer for the first time and now use it frequently, particularly when they are outside their home. However, not all respondents had changed to a proper handwashing technique. Some respondents reported only using water to wash their hands, sometimes due to the cost of soap (Table 2).

**Table 2.** Reported changes and challenges in handwashing practices as a result of COVID-19 or lockdowns.

| Handwashing Practice | Quantitative Results | Qualitative Themes and Descriptions |
| --- | --- | --- |
| Changes related to handwashing | 86% (N=104) of respondents reported coronavirus or the lockdown had changed their handwashing practices<br>72% (N=21) of caregivers reported coronavirus or the lockdown had changed the handwashing practices of young children (<5 years old) in the household | • <u>Increased frequency of handwashing:</u> Among respondents who reported a change, the majority said they were washing their hands more frequently now due to COVID-19. Some respondents described washing hands very frequently, such as 20 to 30 times a day or every 30 to 60 minutes. Many explained they are now more conscious or alert about handwashing and are washing their hands as soon as they get home, after each task, or after touching objects outside the home, as well as at key times such as before eating. Some respondents discussed how the pandemic had made handwashing a habit for them. Others specifically said that no one washed their hands before coronavirus and it was a new practice for them.<br>• <u>Started using any or more soap/sanitizer:</u> Many respondents explained they started using soap or waterless hand sanitizer due to COVID-19 or reported using more soap or water now. Several respondents also elaborated that this is their first time using hand sanitizer and they now keep a bottle with them and use it frequently.<br>• <u>Started washing hands more thoroughly:</u> Some respondents explained that they now wash their hands more thoroughly. This included washing hands for a longer time and washing specific parts of the hands, such as the fingers and gap between the fingers that they had not previously washed. A few respondents said they had changed their practices after learning about proper handwashing practices from ASHA and Anganwadi workers and one respondent explained he had learned the technique for washing his whole hands from television.<br>• <u>No change:</u> Among respondents who reported no change in their handwashing practices, many said this was because they had always washed their hands with soap and water often.<br>• <u>Improper handwashing:</u> Not all respondents had changed to a proper handwashing technique. Some respondents reported only using water to wash their hands. For some, this was due to the cost of soap, although not all respondents explained why they only use water. A few respondents also said they do not wash their hands frequently because they are not used to it or because they only wash their hands when they remember. |
| Challenges related to handwashing | 13% (N=17) of respondents reported challenges to frequently washing their hands with water and soap | • <u>Challenge buying soap (8% of all respondents, N=10):</u> The challenge with soap was predominantly cost, with two respondents specifically stating the government had not offered help with buying soap. No one mentioned being unable to find soap at shops.<br>• <u>Challenge getting water (6% of all respondents, N=8):</u> For challenges with water, respondents described problems discussed in more detail in Table 3 under "Challenges related to household water availability".<br>• <u>Other challenges (2% of all respondents, N=2):</u> One respondent described how frequent handwashing can negatively impact your skin and another said that it can be challenging to use soap regularly as it is a new practice. |

When describing their increase in handwashing frequency and soap/sanitizer use, many respondents explained that they were now washing their hands very frequently, with several respondents linking handwashing to new daily activities or discussing that it has become a new habit, whereas they previously only washed before eating or not at all:

> *“Earlier, we did not use to wash our hands when we used to come from outside. We used to wash our hands only when needed. Now, when we come back from outside or market, we wash our hands and feet before entering the house. We are washing our hands 5-6 times a day. If we have to go to some office, we wash our hands. We are using [waterless hand] sanitizers as well. I wash my hands and use the sanitizer afterwards at times. The quantity of soap and water has also been increased*.
>
> *” - Male respondent, 35-39 years old (June 2020)*
>
> *“Handwashing has turned out to be a habit. Everyone is doing it. I never used to wash my hands, but after coronavirus, we have been washing them frequently*.
>
> *” - Male respondent, 45-49 years old (July 2020)*
>
> *“We are cleaning our hands as said in the TV. Earlier, did we use to wash our hands 20-30 times with soap in a day [like we are doing now]? We used to only wash once or twice [a day] before having food*.*” -Male respondent, 55-59 years old (May 2020)*
>
> *“We did not use to wash our hands earlier, but are now washing them with soap and sanitizer. In every 20-30 minutes, we wash. Who used to wash their hands like this earlier, madam? They used to wash, but only before eating*.
>
> *” - Female caregiver respondent, 25-29 years old (July 2020)*

Some respondents also explained that they had made changes to their handwashing practices to now wash their hands more thoroughly:

> *“Now, after seeing it from the TV, we are washing our hands very frequently from all the sides in a systematic way; from above, below and between the fingers. Earlier we only used to wash one side of the hand palm and that too, not very frequently*.
>
> *” - Male respondent, 65-69 years old (June 2020)*

Caregivers were also asked if the handwashing practices of their young children (<5 years old) had changed as a result of COVID-19 or the lockdown, to which 72% (N=21) reported ‘yes’. Many of the changes in children’s handwashing practices mirrored the changes in adult’s handwashing, including caregivers washing their young children’s hands more frequently or asking older children to wash their own hands more frequently, and starting to use soap or sanitizer to clean children’s hands.

> *“The children are made to wash their hands every time. For example, if they are playing inside the home or studying, after that, their hands are washed. Their hands are washed some 10-12 times in a day. We never used to do it earlier, but have started doing it only after coronavirus*.
>
> *” - Female caregiver respondent, 25-29 years old (June 2020)*

### Challenges related to handwashing

Most respondents (87%, N=110) reported no challenges in being able to frequently wash their hands with water and soap. Among the few who experienced challenges (13%, N=17), this mostly related to buying soap (8% of all respondents, N=10) and/or getting water (6% of all respondents, N=8; 3 of these respondents described a challenge with both), with 2 respondents (2%) describing other challenges. The challenge with soap was predominantly cost, with two respondents specifically stating the government had not offered help with buying soap. No one mentioned being unable to find soap at shops. For challenges getting water, this was often due to water scarcity in the summer season or the water source being far away (Table 2).

### Water access and treatment

At the village level, about half of VWSC members (52%, N=11) reported a change to the piped water supply service in their village as a result of COVID-19. Changes included adding bleaching powder to the water tank for water treatment, cleaning the water tank, and/or extending or reducing the hours of water supply (Table 3). At the household level, 84% of respondents (N=101) reported they had not experienced any reduced water availability due to COVID-19 or the lockdown in the past 7 days. Among the few respondents who reported reduced water availability (16%, N=19), many of the problems described were likely unrelated to COVID-19 (Table 3).

**Table 3.** Reported changes and challenges in water access and treatment practices as a result of COVID-19 or lockdowns.

| Water access and treatment | Quantitative Results | Qualitative Themes and Descriptions |
| --- | --- | --- |
| Changes related to village piped water | 52% (N=11) of VWSC members reported a change had been made to their village piped water supply service due to COVID-19 | • <u>Added bleaching powder to village water tank (33%, N=7)</u>: The most common reported change was that bleaching powder had been added to the water tank by the VWSC or government officials, either specifically or more frequently due to COVID-19.<br>• <u>Cleaned village water tank (24%, N=5)</u>: The next most common reported change was the water tank being cleaned by the VWSC or government officials, either specifically or more frequently due to COVID-19. As an example, one VWSC respondent said they used to clean the tank regularly once every two months, but now increased the cleaning frequency to once per month due to COVID-19.<br>• <u>Change in water supply service hours (14%, N=3)</u>: In two villages, the water supply hours were *extended* due to coronavirus. In one of these villages, the VWSC member said they extended the water supply time each day due to the water needs of people staying at the village quarantine center. In contrast, in another village the water supply hours were *reduced* due to its village quarantine center using a lot of water.<br>• <u>No change (48%, N=10)</u>: Among the other half of VWSC respondents who reported no change in the piped water supply service due to coronavirus, four explained they already regularly clean the tank and/or put bleaching powder in it, and three explained the water is already supplied throughout the entire day. Four villages also mentioned issues related to water scarcity and the water source for the tank drying up in the summer season. |
| Changes related to household drinking water practices | 17% (N=20) of respondents reported that coronavirus or the lockdown had changed their drinking water practices | • <u>Started treating their drinking water (16%, N=19)</u>: The following changes in drinking water treatment due to COVID-19 were reported: 16 respondents started to boil their drinking water, 1 started to both filter and boil their water, 1 treated their household water tank with bleaching powder, and 1 treated their water with a tablet that was given to them by an ASHA worker. Several respondents that reported no change in their drinking water practices explained they do not treat their drinking water in any way, although many also reported they have always treated their drinking water, either by boiling, filtering, or using an Aquaguard purifier attached to their tap.<br>• <u>Changed how they store their drinking water (3%, N=4)</u>: Four respondents explained that they either cleaned or changed their drinking water storage container due to coronavirus. Specifically, one respondent said they cleaned their water storage containers now, another said they were using a cleaner vessel, one started storing water in clay pots that were distributed to them, and lastly, one respondent had stopped putting their hands in the water container when getting water out and were instead using a utensil to retrieve the water. |
| Challenges related to household water availability | 16% (N=19) of respondents reported they had experienced reduced water | • <u>Reduced water availability due to COVID-19</u>: Only two respondents provided details of water availability problems specifically related to coronavirus. One respondent used to get water from public taps including one at the school, but due to the lockdown they can no longer get water from the school tap, making it more difficult for them to |
|  | availability in the past 7 days | access water. Another respondent explained they are getting less water now because people residing at the village quarantine center are using water, which has led to fewer hours of piped water supply.<br>• <u>Reduced water availability that may be partially related to COVID-19:</u> Respondents in four different villages described recent problems with piped water availability. Three of these respondents explained that the motor for their village's water tank pump was not functioning and had to be repaired, but it was fixed now. It was not mentioned whether COVID-19 impacted the ability to fix the pump quickly, but it is possible this was a factor. In one village the respondent said the motor was not working for 20 days.<br>• <u>Reduced water availability not related to COVID-19:</u> Several respondents explained problems with water availability that were not related to COVID-19. At least four respondents described water scarcity being a problem in the summer season, due to water sources drying up, while a couple respondents said the piped water becomes muddy when it rains. Others mentioned general problems with the water supply system, such as difficulty with the water connection, problems due to an old pipeline or changes to the water supply system, not getting regular water, or that water is only supplied for 30 minutes a day. |

The majority of respondents 83% (N=98) reported no change in their drinking water practices due to COVID-19. Among the respondents who reported a change (17%, N=20), 19 respondents (16% of total respondents) described a change in how they treat their drinking water, and 4 respondents (3% of total respondents) described a change in how they store their drinking water (3 of these respondents described a change in both; Table 3).

### Sanitation

The pandemic and lockdown had no major impact on reported latrine use (Table 4). Almost all respondents (96%, N=109) reported no change in their family’s defecation practices as a result of COVID-19 or the lockdown, and 88% (N=100) reported defecating in the latrine with only 12% (N=14) defecating in the open. Similarly, no caregivers reported a change in their child feces management or latrine training practices due to COVID-19 or the lockdowns, although some described impacts on general childcare practices and/or support (Table 4). Among the few respondents who did report a change of practices in their family, one respondent reported a change in his grandfather’s sanitation practices due to fear of the virus:

**Table 4.** Reported changes and challenges in sanitation practices as a result of COVID-19 or lockdowns.

| Sanitation Practice | Quantitative Results | Qualitative Themes and Descriptions |
| --- | --- | --- |
| Changes related to defecation locations and latrine use | 88% (N=100) of respondents reported defecating in the latrine the last time they defecated<br>3% (N=3) reported a change in their family's defecation practices due to coronavirus or the lockdown<br>9% (N=10) thought there was more latrine use in their village now | • <u>Small change in household latrine use:</u> Among the few respondents who reported a change in latrine use practices, one respondent explained that their grandfather became scared of defecating in the open due to the COVID-19 outbreak and started defecating in the latrine. Two other respondents talked about specifically using phenyl or "medicine" to clean the latrine after defecation due to coronavirus.<br>• <u>Small increase in village latrine use:</u> When considering defecation practices in their entire village, only 9% (N=10) believed there was more latrine use in their village now. Among those who reported more latrine use in their village, some respondents explained that this change was due to the increased awareness or fear of coronavirus, whereas others said that villagers are going out less now due to the lockdown. More specifically, a VWSC member explained when villagers used to leave the village to go somewhere such as marketplaces, they would defecate in the open due to unavailability of a latrine there. However, since that movement has stopped, that open defecation has also stopped. Among those that reported no change in village defecation practices (77%, N=86), many reported that everyone in the village was already using the latrine before coronavirus. For respondents who said they did not know if there was a change (14%, N=16), these were mostly caregivers who explained that they do not know other villager's defecation practices.<br>• <u>No change in household latrine use:</u> Among respondents that reported no change in their family's latrine use, most elaborated that all family members always use the latrine (83%, N=91), which was also the case before COVID-19. Some respondents specifically mentioned they have been using the latrine for a long time. Other respondents (10%, N=11) reported that they sometimes open defecate and sometimes use the latrine. Some of these respondents explained that the decision of whether or not to use the latrine was based on time of day (e.g., open defecate in the daytime but use the latrine at night due to snakes), water supply (e.g., go for open defecation when there are problems getting water or water supply has stopped), season (e.g., prefer to use the latrine during rainy season), or condition of the latrine (e.g., do not use latrine when wall or latrine pan are broken). A few respondents (5%, N=6) explained that some family members open defecate while others use the latrine. Out of the four respondents who said they defecated in the open, three did so because they did not have a latrine while one explained this is a habit he cannot change. |
| Changes related to child feces management (CFM) | No caregivers reported a change in CFM practices connected to the pandemic/lockdowns | • <u>No change in CFM practice due to COVID-19:</u> Only four caregivers (12%) reported a change in where their child defecates and where they dispose of their child's feces since the start of the pandemic or lockdowns. However, none attributed this change to the pandemic, but rather to their child growing and transitioning to new behaviors: two of the children started walking and hence began learning to use the latrine while the other two children had started their latrine training and then fully transitioned to latrine use. Although there was no change for households interviewed, two caregivers thought that child latrine use must be happening more now in their village as families are doing whatever they can to keep their children healthy and safe from coronavirus. |
|  |  | • <u>Change in childcare and support caregivers receive:</u> About one fourth of caregivers (26%, N=9) described a change in childcare or support with childcare tasks such as CFM. Six caregivers (18%) described being 'more alert' in their childcare, such as paying closer attention to their child's hygiene and ensuring they do not roam outside, two caregivers (6%) received more childcare support now with more family members at home, and one caregiver (3%), in contrast, expressed less support since she could no longer visit relatives. |

> *“Whoever does not have the habit of going to the latrine goes outside. It is my grandfather. Now, after coronavirus, he is not going out anymore. He is getting scared. Rest of us have always been using the latrine*.
>
> *” - Male respondent, 25-29 years old (June 2020)*

Respondents were also asked if COVID-19 or the lockdowns had impacted defecation practices within their village. Again, the majority of respondents (77%, N=86) believed there had been no change, 14% (N=16) did not know whether there had been a change, and only 9% (N=10) believed there was more latrine use in their village now. Among those who reported more latrine use and less open defecation in their village now, it was common for respondents to attribute this change to either awareness of coronavirus or because villagers were going out less due to the lockdown:

> *“Half of the people who believe in coronavirus, use the latrine and bathroom. The ones who do not believe in it go to the open*.
>
> *” - Female respondent, 25-29 years old (July 2020)*
>
> *“Everyone’s latrine is in use since before [coronavirus]. However, the way people used to go outside is not happening anymore. This is not only out of the fear of corona, but also due to lockdown. People also get beaten up if they go out now. [Before the lockdown] when there is a weekly market in another village or town which people attend and feel like defecating at the same time, they have to go out as there is no latrine in the market areas*.
>
> *” - Male respondent, 30-34 years old (July 2020)*

Additionally, although the interview did not include a specific question related to latrine construction, two respondents from households without latrines expressed that COVID-19 or the lockdown had either stopped or made it more difficult for them to construct a household latrine. One respondent described how his family was previously building a latrine, but it had stopped due to COVID-19. Another respondent described how her family wanted to build a latrine but it was now difficult to do so because of the lockdown and low income due to COVID-19.

### Household cleaning

> Over a third of respondents (41%, N=49) reported that COVID-19 or the lockdown changed their household cleaning practices (Table 5). All respondents who reported a change described a positive change, including cleaning the house more frequently since the pandemic started (34% N=40) and either starting to use detergent/disinfectant or using more of it now (13%, N=16):
>
> Table 5. Reported changes and challenges in household cleaning practices as a result of COVID-19 or lockdowns. 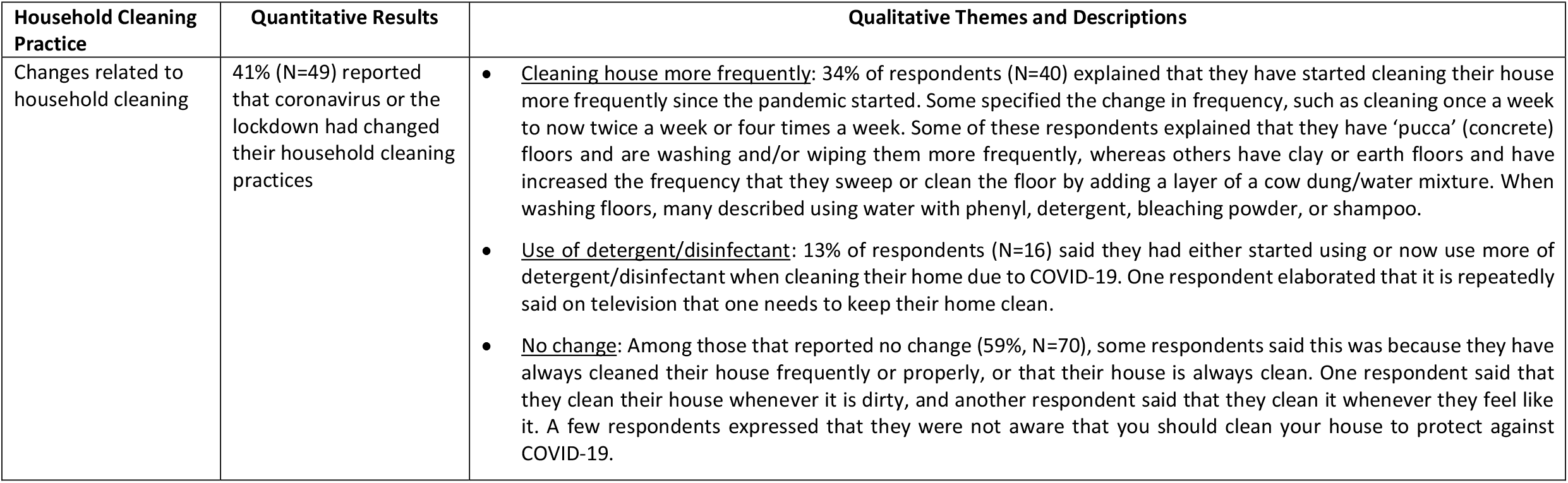
>
> *“When it comes to cleaning our house, we are doing it more than how we used to do it earlier. Now, after coronavirus, instead of cleaning the house once a week, we have to do it four times. After washing, we have to sanitize it and put some phenyl and bleaching powder*.
>
> *” - Male respondent, 65-69 years old (June 2020)*
>
> *“We are staying clean. Earlier, we may or may not have washed the house, but now we have to wash it*.
>
> *” - Female caregiver respondent, 30-34 years old (June 2020)*
>
> *“Yes, it has been communicated by TV that we should keep our house clean. People are also getting aware of this. Now, there is no work for me. I have to spend all my time in cleaning the house now. As I do not have to do any work outside my home, I have to do some household work*.
>
> *” - Male respondent, 45-49 years old (July 2020)*

## Discussion

The COVID-19 pandemic led to positive changes in WASH practices in rural Odisha, India. In particular, we found the majority of respondents reported increased frequency and/or improved handwashing practices, an important WASH behavior to reduce the spread of COVID-19. We also found improvements in other WASH behaviors as a result of the pandemic for some villages and respondents, including improved service delivery of village level water supply, increased household water treatment, and improved household cleaning practices. There was minimal change in defecation practices as the vast majority of respondents continued to use their household’s latrine. The WASH infrastructure investments in study villages, including village-level piped water supply and high household latrine coverage, likely aided in compliance with COVID-19 preventative measures – particularly the ability to wash hands frequently and stay home during strict lockdown periods. These findings can be useful for improving response to COVID-19, understanding the effects of the pandemic on WASH behavior change and habit formation, and building resiliency to be better prepared for future pandemics.

Participants overwhelmingly reported positive changes in handwashing due to the pandemic (with 86% reporting a change), and few reporting challenges with frequently washing their hands with soap and water. We found an increase in handwashing frequency and thoroughness, as well as the use of soap, which remained in place a few months after the pandemic began and the first lockdown went into effect in India, suggesting a new habit may have formed among adults and children. Many respondents also linked handwashing behavior as a new and consistent practice after additional actions throughout the day (such as returning home), offering further evidence that habit formation was occurring for handwashing, which is an important predictor for achieving sustained handwashing behavior change.^15,16^ The longevity of these changes also suggests that improved handwashing habits might remain after the pandemic subsides, which could potentially lead to reduced disease transmission for other respiratory and fecal-oral diseases, as was observed in Mexico after the H1N1 influenza pandemic.^17^ Among the few participants that reported barriers to handwashing in our study, this was attributed to the cost of soap (8%) or challenges getting water (6%) that were often due to a problem with the water supply system or water scarcity in the summer season. The proportion of respondents reporting barriers to handwashing was substantially lower than a study among students and slum dwellers in Uganda where 60.1% reported lack of soap, detergents, alcohol-based hand rub, or antiseptic as a barrier to handwashing and 33.9% reported lack of running water as a barrier.^18^ These findings illustrate how the provision of soap and reliable water supply are needed to enable individuals to practice the promoted preventative measure of handwashing. This is also in line with common behavior change theories which include the concept of having an enabling physical environment as a requisite for behavioral performance.^19–21^ Therefore, the provision or facilitation for accessibility of infrastructure along with mobilization towards proper use may be required to achieve behavior change for many WASH practices and preventative measures.

Our results related to handwashing as a COVID-19 preventative measure are generally in agreement with other studies. There were high levels of reported handwashing as a preventative measure in online surveys throughout India,^22–26^ a phone survey in Tamil Nadu, India,^27^ and in-person surveys in Ethiopia^28^ and Kenya,^29^ all of which found at least 85% of respondents reported handwashing to prevent COVID-19. However, many of these surveys did not specify water source access, the use of soap, or if there was a change in handwashing practices.

Additionally, most of the online surveys were in English and required literacy and internet access, which likely skewed the population to wealthier urban respondents not representative of rural villagers. Furthermore, the wording of the survey question could impact response, as seen in our survey where 86% reported that COVID-19 or the lockdown had changed their handwashing practices for the better, compared to 96% who reported that they had washed hands/used hand sanitizer more frequently in the past 7 days as an action to avoid COVID-19.^11^ This difference is likely due to the more qualitative nature of the former question that prompted respondents to explain changes and give more detailed responses about their handwashing practices. Overall, this study population had high levels of knowledge about the importance of handwashing and other preventative measures to reduce transmission of COVID-19, and reported primarily getting their information about COVID-19 from television news, conversations with a community-level government worker, and/or social media or the internet.^11^

Some households described improvements in other healthful WASH behaviors, such as water treatment and/or household cleaning practices, which could also have beneficial effects on overall disease transmission. Although the majority of households in these villages have functional piped water connected to their households, the supply is intermittent in many cases, often with set daily hours of operation, and many households are still storing water as a result.^30^ Therefore, point-of-use treatment of water may still be required to ensure safe quality drinking water in many of the villages where regular chlorination is not provided. The reported water treatment and household cleaning behaviors may have been a kind of spillover behavior change effect resulting from other messaging on preventing COVID-19 transmission. In addition, VWSC members in many villages reported that government officials came to clean village water tanks or add bleaching powder to the tank for water treatment, and these actions may have given focus to proper water treatment and encouraged a general message of cleanliness. While these WASH behaviors have not yet been proven to reduce transmission of COVID-19, they could reduce transmission of other diseases such as diarrheal disease.^31^

There was minimal change in defecation location as a result of the lockdowns or COVID-19, with most respondents reporting continued latrine use. This is in line with the results of another study in Tamil Nadu, India, which also found minimal change in defecation practices with 92% of respondents reporting no change since the lockdown and the majority of respondents with a private latrine continuing to use it and only a few (7%) beginning to use their private latrine as a result of the pandemic.^27^ Our study area had high levels of latrine coverage and use prior to the pandemic,^30^ which may have also contributed to the minimal change observed. Our qualitative findings also suggest little change in the decision-making or motivational factors individuals typically consider when choosing their place of defecation. Many respondents who reported no change described aspects such as time of day, water supply, season, or condition of the latrine as driving their defecation practice, which aligns with factors noted in previous work completed prior to the pandemic in the nearby district of Puri in Odisha.^32^ When defecation changes were described by our participants, they were often a result of other factors caused by the lockdown, such as reduced traveling to markets or other public areas that do not have latrines, with few respondents associating defecation practices with COVID-19 risk. Additionally, although there is room for improvement in child feces management practices in this study area,^33^ caregivers did not report any changes in these practices as a result of the pandemic.

Access to adequate WASH infrastructure can play a role in an individual’s ability to comply with recommended COVID-19 preventative measures in two ways. First, adequate handwashing requires sufficient water availability from easily accessible and reliable sources, with household piped water being the highest level of access.^34^ Second, physical isolation at home for individuals suspected to have COVID-19 as well as the general population during periods of strict lockdown have been promoted as measures to reduce transmission. However, these measures are only feasible if there is adequate water, hygiene, and latrine facilities in a household; otherwise household members would need to access public locations to retrieve water, bathe, and defecate.^34^ We found in our study that good WASH infrastructure enabled compliance with preventive measures like handwashing or staying home, as participants were able to use their latrine and wash their hands at home. A separate study in Tamil Nadu, India found that respondents of the same age, gender, and education who had access to a private toilet were more likely to report they increased the frequency of handwashing since the lockdown,^27^ further suggesting that infrastructure played a role in COVID-19 handwashing practices. Overall, these findings suggest that investments in WASH infrastructure, such as a piped water supply, not only ensure individuals have an enabling environment to perform promoted WASH practices but may also play a role in building the resilience of rural communities against future disease outbreaks.

There are some limitations of this study. First, it relied on phone interviews, so we could only include participants with a mobile phone and network connection. This may exclude some of the poorest and most remote households, although we were able to interview some participants who lived in villages without a mobile network by calling them when they were in an area outside of their village that had network. Additionally, responses were self-reported. This could introduce reporting bias, as respondents sometimes overreport hygienic behaviors like handwashing due to courtesy bias or social desirability bias.^35–37^ Self-reports of increased handwashing also do not measure if handwashing is being performed correctly. While some respondents in our study explained they had learned the correct handwashing technique due to COVID-19 information campaigns and are now washing all parts of their hands, this may not be true of the entire study population. For example, in a study in Nigeria, only 39% of respondents washed all critical parts of their hands correctly when asked to demonstrate handwashing, compared to 90.5% of respondents who reported practicing regular handwashing with soap and water to prevent COVID-19.^38^ We tried to reduce reporting bias and capture detailed experiences by including several open-ended questions and asking for follow-up explanations to closed-ended questions. Finally, we targeted respondents who resided in villages that had completed the MANTRA program that installed village-level piped water and created high levels of sanitation access,^30^ and therefore results related to WASH practices may not be generalizable to other villages with lower levels of water and sanitation access.

The research revealed rich descriptions of changes in WASH practices among rural villagers in Odisha as a result of the COVID-19 pandemic, including improvements in handwashing practices that were promoted for COVID-19 prevention, as well as improvements in other WASH practices that were not directly promoted such as water treatment and household cleaning. With regards to handwashing, we found an increase in handwashing frequency, thoroughness, and use of soap, and that these practices remained in place a few months after the pandemic began and were often described as a new and consistent practice after additional daily actions, suggesting new habit formation that could potentially lead to sustained handwashing behavior change. The role of barriers and enabling factors were also described for WASH-related preventive practices, including the provision of soap and a reliable water supply. The results also highlight the importance of adequate WASH infrastructure, including piped water, in enabling resilience and allowing villagers to practice safe behaviors during a large-scale health crisis.

## Data Availability

All relevant data are included within the manuscript. Further inquiries related to data access can be sent to Valerie Bauza.

## Acknowledgments

The authors thank Varsha Priyadarshini and Dhiren Swain for their help with data collection and Sabrina Haque, Sheela Sinharoy, and Miles Kirby for their input related to the interview tool and mobile phone data collection plans. This research was supported in part by grants from the Bill&Melinda Gates Foundation (OPP1125067) and National Institute of Environmental Health Sciences, USA (T32ES012870 to VB).

